# Applications of qualitative grounded theory methodology to investigate hearing loss: Protocol for a qualitative systematic review

**DOI:** 10.1101/19004259

**Authors:** Yasmin H K Ali, Nicola Wright, David Charnock, Helen Henshaw, Melanie A Ferguson, Derek J Hoare

## Abstract

**Introduction:** Hearing loss is a chronic condition affecting 11 million individuals in the UK. People with hearing loss regularly experience difficulties interacting in everyday conversations. These difficulties in communication can result in a person with hearing loss withdrawing from social situations and becoming isolated. While hearing health loss research has largely deployed quantitative methods to investigate various aspects of the condition, qualitative research is becoming more widespread. Grounded theory is a specific qualitative methodology that has been used to establish novel theories on the experiences of living with hearing loss.

**Method and analysis:** The aim of this systematic review is to establish how grounded theory has been applied to investigate the psychosocial aspects of hearing loss. Methods are reported according to the Preferred Reporting Items for Systematic Reviews and Meta-analysis Protocols (PRISMA-P) 2015 checklist. Studies included in this review will have applied grounded theory methodology. For a study to be included, it can apply grounded theory as an overarching methodology, or have grounded theory methodology embedded amongst other methodologies. These studies can be in the form of retrospective or prospective studies, before and after comparison studies, RCTs, non-RCTs, cohort studies, prospective observational studies, case-control studies, cross-sectional studies, longitudinal studies, and mixed method studies. Purely quantitative studies, studies that have not applied grounded theory methodology, articles reporting expert opinions, case reports, practice guidelines, case series, conference abstracts, and book chapters will be excluded. Studies included will have adult participants (≥18 years) who are either people with an acquired hearing loss, their family and friends (communication partners), or audiologists. The quality of application of grounded theory in each study will be assessed using the Guideline for Reporting and Evaluating Grounded Theory Research Studies (GUREGT).

**Ethics and dissemination:** As only secondary data will be used in this systematic review, ethical approval is not required. No other ethical issues are foreseen. The International Prospective Register of Systematic Reviews (http://www.crd.york.ac.uk/PROSPERO) holds the registration record of this systematic review. Findings will be disseminated via peer reviewed publications and at relevant academic conferences. Findings may also be published in relevant professional and third sector newsletters and magazines as appropriate. Data will inform future research and guideline development.

**Prospero registration number:** PROSPERO CRD42019134197

**Strengths and limitations of this study:**

- This systematic review is the first to provide a comprehensive critique of the use of grounded theory to investigate hearing loss.
- The search strategy was formed in collaboration with an information specialist at the University of Nottingham.
- The PRISMA-P guidelines have directed the considerations and layout of this protocol.
- Because experiences and articulations of hearing loss are influenced by age, only adult (≥18 years) participants (people with hearing loss, communication partners, audiologists) will be considered.
- The search will not include grey literature.
- The studies included will only have samples of individuals with hearing loss, rather than full deafness.

## INTRODUCTION

Hearing loss is a chronic condition that involves a decrease in an individual’s ability to hear sounds. It can occur at mild, moderate, severe, or profound levels, and typically worsens over time^1^. Eleven million people in the UK currently live with hearing loss ^2,3^. This number is expected to increase to 15.6 million by 2035^3^. Globally, over 900 million people are expected to acquire a disabling hearing loss by 2050 ^2,3^.

The annual global societal cost of unaddressed hearing loss is $750 billion^4^. Within the UK, the annual cost of hearing loss is £30 billion, with the larger proportion of this cost dealing with the social, psychological, and health impacts of hearing loss, rather than the treatments provided by audiology services^5^. Untreated hearing loss can impact the work opportunities and occupational progression of people with hearing loss (PHL)^4^. Estimations of the global occupational limitations occurring due to the stigma and inequalities associated with hearing loss is $105 billion annually^4^.

PHL experience functional limitations such as communication difficulties in day to day conversations, where speech is often misheard and becomes challenging to follow^6-8^. Failed instances of communication can lead PHL to experience embarrassment, e.g. if incorrect responses are given in a conversation after mishearing others^9,10^. Because PHL struggle to have full interactions with others they can come to feel isolated and separated from the world^10-12^. The anxiousness of repeatedly not being able to hear others can lead to withdrawal from social situations^13^. Consequently, it is estimated that PHL are more likely to develop depression than the general population^14-17^.

The impact of hearing loss is not only psychological, as it can also impact interpersonal relationships of PHL also. Not being able to fully communicate and establish successful interactions can also lead to feelings of frustration^18^. Individuals that communicate with PHL, such as their spouses, family and friends, can also become frustrated due to not being heard or understood in interactions^18,19^. Conflict in the relationships of PHL and their communication partners have been reported to occur^18,20,21^, with hearing loss being a significant risk factor for the development of abusive relationships^22^.

Investigations into the experience of hearing loss are crucial to understanding the implications it has for the person, and the implications it has for their care. To date, most studies in hearing loss research have used quantitative methods. Due to the evident psychosocial impacts of hearing loss, there has been a significant increase in the adoption of qualitative methodologies in hearing loss research^23^, particularly since the publication of the “*Conducting qualitative research in audiology: A tutorial”* in 2012^24^. Qualitative methods allow researchers to understand the experiences, opinions and perspectives associated with hearing loss, which experimental measures are not designed to uncover^23-25^. Five main qualitative methodologies exist: phenomenology, ethnography, case studies, narratives, and grounded theory^26^. Each of these methodologies aims to uncover the experiences of a particular sample in different ways, depending on the type of knowledge to be established^26^. Grounded theory is the only qualitative methodology that is exclusively used to establish novel theories completely uninfluenced by previous knowledge^26^. Grounded theory was first established by Glaser and Strauss in 1967^27^. It is defined as the systematic exploration of data in an open-minded, comparative, and rigorous manner for developing a novel theory that is purely grounded within the data^28,29^. Through applying grounded theory, people’s experiences, and occurring social processes can be understood by integrating definitions and meanings from the perspectives of individuals from the target population^30,31^. To do this, a researcher can adopt the principles of one of three grounded theory schools (Table 1).

**Table 1.** The three different schools of grounded theory methodology

| Dimensions of Comparison | Glaserian School (1967)<sup>27,29</sup> | Straussain School (1990)<sup>31</sup> | Constructivist School (2006)<sup>28</sup> |
| --- | --- | --- | --- |
| Philosophical stance | Empiricism | Interpretivism | Constructivism |
| School founders | Barney Glaser & Anselm Strauss | Anselm Strauss & Juliet Corbin | Kathy Charmaz |
| Philosophical principle | Knowledge is formed based on individual experience, and can be objectively measured. | Knowledge is subjective and socially constructed. Establishing one 'truth' is impossible. | Knowledge is subjective and is constructed by both the participant's experiences and the researcher's own interpretations. |
| Researcher's influence | Researchers can completely detach themselves from their research and not influence it. | Researchers can't detach themselves from the research and will always influence the research process and findings. | A comprehensive truth is pursued, however it will only be reflective of the social context and group being studied. |
| Grounded theory emphasis | *Constant comparative analysis*: constantly comparing data and outcomes to establish objective knowledge. | *Reflexivity*: researcher provides reflections on the process of data collection, analysis, and recognises how they influence this process. | *Constant comparative analysis, reflexivity, theoretical sampling* (knowledge is pursued and collected through recruiting different samples and investigating different concepts to develop the theory). |

Assessing the methodological applications of grounded theory is essential for determining the trustworthiness and credibility of the theories developed, as suggested by researchers in various fields such as psychology^32^, nursing^33^; physiology^34,35^ and business & management^36^. A field that has applied and critiqued applications of grounded theory is chronic illness research^37-40^. The founders of all three schools of grounded theory (Table 1) also specialise in investigating chronic illnesses. Glaser & Strauss initially formed the methodology to investigate fatal chronic illnesses^27,41^, but then used it more extensively to investigate non-fatal chronic conditions^38,42,43^. Charmaz has since identified the methodology as the most appropriate for establishing novel knowledge regarding life-long conditions^28,38,43-45^.

Hearing loss is also a chronic disease as recognised by the World Health Organization^46,47^, and is the third most common chronic condition affecting the population^48-50^. Despite this none of the aforementioned research into chronic illness reviewing grounded theory included hearing loss as a condition for consideration. Systematic reviews that have investigated hearing loss to date have mainly focused on reviewing phenomena related to hearing loss, such as its impact on interpersonal relationships^18^, hearing aids and hearing loss^51^, hearing loss and alternative listening devices^52^, treatments for PHL with cognitive impairments^53^, hearing loss and auditory training^54^, vertigo and hearing loss^55^, quality of life and hearing loss^56^, and hearing loss and depression^57^.

No systematic review investigating hearing loss has yet assessed the use of qualitative methodologies to reach these findings, including grounded theory. The current systematic review will be the first to systematically examine the application of grounded theory methodology to investigate hearing loss. The review aims to (1) describe how grounded theory methodology has been applied within the field of hearing loss, (2) critically appraise the execution of grounded theory methodology in each study, and (3) produce recommendations for future grounded theory methodological applications related to hearing loss.

## METHODS AND ANALYSIS

The methods and analysis of this systematic review will be reported in accordance to the Preferred Reporting Items for Systematic Reviews and Meta-Analysis (PRISMA) 2015 checklist^58,59^. The recommended items on the PRISMA-P checklist will form the subheadings of this section. This systematic review will also follow the Cochrane Handbook’s suggested approach for conducting methodology-based systematic reviews, i.e. SDMO structure (Studies, Data, Methods, and Outcomes) ^60,61^. The SDMO is used to investigate contemporary research methods to illuminate the impact of the methodology on the quality of research within a specific field^60^.

## ELIGIBILITY CRITERIA

### Types of Studies

Studies included in this review will have applied grounded theory methodology. For a study to be included, it can apply grounded theory as an overarching methodology, or be embedded into it amongst other methodologies. These studies can be in the form of retrospective or prospective studies, before and after comparison studies, RCTs, non-RCTs, cohort studies, prospective observational studies, case-control studies, cross-sectional studies, longitudinal studies, or mixed method studies. Purely quantitative studies, studies that have not applied grounded theory methodology, articles reporting expert opinions, case reports, practice guidelines, case series, conference abstracts, and book chapters will be excluded.

### Types of Data

Original research submitted to peer-reviewed journals will be the data used in this systematic review. The data collected in those included studies will include qualitative primary data occurring in the form of transcripts or quotes from audio interviews with participants, patient journals, written reflections of patients, family members of PHL, or audiologists, memos of progression of study, initial themes and analysis, and observational. Records reporting diary entries of patients before participation in their prospective study, will also be eligible for inclusion. No secondary data will be collected.

### Types of Methods

The methodology under review in this systematic review is qualitative grounded theory methodology. Different methods such as interviews, observations, and focus groups, can all be undertaken using grounded theory methodology. Based on the principles of grounded theory, and the school of grounded theory followed, the most appropriate methods are selected. The methods used in each study will be identified and discussed.

### Types of Outcomes

The explicit use of grounded theory methodology as outlined in the methods section of studies investigating hearing loss. Where available, specific mentions of the applications of grounded theory and the type of school followed will be extracted. Studies must also have researched topics regarding hearing loss with grounded theory methodology, and these topics will also be extracted and collated.

### Settings

Any research setting will be included.

### Participants

Participants will include PHL with some residual hearing. Studies involving deaf individuals with no residual hearing left will be excluded. This is because being deaf entails significant different experiences to having a hearing loss. Most deaf individuals incorporate being deaf as part of their identity, and learn sign language at an early stage and become a part of the deaf community^62,63^. People with hearing loss on the other hand view their condition as a disability which limits them in their everyday lives, and are less likely to use sign language or be a part of the deaf community as they do not share that identity^64^.

Only studies including adults (≥18 years) will be included. The levels of hearing loss for eligibility are mild, moderate, severe, or profound, according to the British Society of Audiology (2018)^65^. Cochlear implant users, hearing aid users, and non-hearing aid users will be included. Studies involving communication partners (friends, family members, and colleagues that interact with PHL) will also be eligible for inclusion. Finally, studies involving audiologists that are involved in hearing loss treatment will be included. Studies with children and young adolescents (<18 years) will be excluded, unless they are in a study with adults where the adult data is reported separately.

### Information sources

A systematic search strategy will be employed to identify peer-reviewed journal articles that meet the eligibility criteria. The following databases will be used: PsycINFO (1800S- current), ASSIA (Applied Social Sciences Index and Abstracts, 1987 – current) Global Health (OvidSP database, 1973- current). Web of Science (1899- current) Pubmed (1996- current) British Nursing index (1994- current) CINAHL (Culmative Index to Nursing and Allied Health Literature, 1961- current) MEDLINE (Ovid, In-process & Other Non-Indexed Citations, 1946- current) Scopus (1983- current) EBSCO (1944- current). Google Scholar will be used for forward citation tracking.

Additionally, snowballing of the reference lists of articles shortlisted for inclusion will be undertaken, and related articles from shortlisted authors will be screened in an attempt to identify any relevant articles which may not have been returned by the database searches. The electronic database searches will be updated just before the final analyses and any eligible studies retrieved will be included. At the end of the study selection process, the search strategy produced for each database will be reported, in addition to a PRISMA flow diagram.

### Search Strategy

The search strategy was formed from free text and controlled vocabularies (Medical Subject Headings (MeSH)), consultations with an information specialist at the University of Nottingham, and the testing of search results.

1. Hearing Loss, Noise-Induced/ or Hearing Loss/ or Hearing Loss, Sensorineural/ or Hearing Loss, Unilateral/ or Hearing Disorders/ or Hearing Loss, Sudden/ or Hearing Loss, Bilateral/ or Hearing Tests/ or Hearing Loss, Conductive/
2. hearing loss*.mp. or exp Hearing Loss/
3. 1 or 2
4. Amplifiers, Electronic/ or amplifier*.mp.
5. Hearing Aids/ or listening device*.mp.
6. Cochlear Implants/ or cochlear implant*.mp. or Cochlear Implantation/
7. exp Hearing Aids/ or hearing aid*.mp.
8. 4 or 5 or 6 or 7
9. hard of hearing*.mp.
10. communication partner*.mp.
11. audiologist*.mp. or Audiologists/ or Audiology/
12. exp Persons With Hearing Impairments/
13. 9 or 10 or 11 or 12
14. grounded theory*.mp.
15. exp Grounded Theor*/
16. 14 or 15
17. 3 or 8 or 13
18. 16 and 17

### Data management

YA will be responsible for management of the data. A digital record will be kept for all identified articles, and the process of data screening and extraction will be tracked in Covidence. A unique ID code will be assigned to the included articles for access to the full text and data collection sheet.

### Selection Process

A review of all titles and abstract resulting from the searches will be performed by YA. During this screening stage, 20% of titles/abstracts will be independently reviewed by a second author. Any differences will be resolved by a third author. Full text screening will be carried out by YA and 20% of full texts will be independently screened by a second author. Any differences between the two authors will be resolved by a third author. Studies that do not meet the inclusion criteria will be excluded from the review and the reason for their exclusion at full text stage will be reported.

### Data extraction process

Extracted data will be in the form of direct text collected from the included articles. Data will be stored in Covidence. Prior to starting data extraction guidance notes will be formed by YA.

Extracted data will include:

1. The study’s title, authors, date of publication, number of citations and country;
2. Date(s)/time period of data collection;
3. Aims, objectives, and/or research questions;
4. Participant characteristics: e.g. age, sample size, gender, country and occupation status;
5. Key participant characteristics: type and severity of hearing loss, hearing aid usage, and whether they have a cochlear implantation;
6. Type of participants included (PHL, their family/ friends - communication partners, audiologists, healthcare practitioners);
7. Methods of recruitment of participants;
8. Data collection methods;
9. Particular type of experience of hearing loss being investigated;
10. School of grounded theory followed if mentioned;
11. Ethical standards/approval;
12. Attempts to establish qualitative rigour or trustworthiness;
13. Study/methodology limitations;
14. Advantages and disadvantages of using grounded theory if explicated by authors;
15. Study design: methodology and methods;
16. Key findings;
17. Conclusions;
18. Recommendations.

### Methodological quality of the individual studies

Quality appraisal of studies will be conducted using the Guideline for Reporting and Evaluating Grounded Theory Research Studies (GUREGT)^66^, and critiques of the methodological quality of each individual study will be established.

YA will appraise all records, and 20% of the studies included will be independently appraised by a second author using the same guidelines.

### Data synthesis

A thematic synthesis approach as established by Braun and Clarke will be used to synthesise the findings^67,68^. Through this approach, six steps are undertaken to achieve codes, refine themes, and synthesise the data. The six stages are: data familiarisation, generation of initial codes, searching for themes, reviewing themes, refining and defining themes, and reporting findings. Analysis will firstly consist of coding the data extracted from the included articles and applying initial codes which are close to the analysed data. This will involve establishing which aspects of hearing loss were investigated, how they were investigated, the use of grounded theory, and the grounded theory processes applied within a study. Codes will subsequently be grouped into more focused themes based on the relevance of each concept. These descriptive themes will aid in outlining how grounded theory has been used as a methodology to investigate hearing loss. In the final stage of analysis, analytical codes of the applications of grounded theory and the implications these have for research when investigating hearing loss will be established. Recommendations for how to apply grounded theory when investigating hearing loss will be made.

To ensure inter-rater reliability as required for qualitative rigour^69,70^, 20% of included articles will be analysed by a second author. Comparisons of results will be established through meetings and any disputes will be resolved by a third author.

### Subgroup analysis

If indicated, studies that are rated as having low quality according to the GUREGT tool will be reviewed separately, to establish the issues that should be resolved when applying grounded theory to investigate hearing loss.

### Ethics and dissemination

As only secondary data will be used in this systematic review, no ethical approval is required. No other ethical issues are foreseen. The International Prospective Register of Systematic Reviews (http://www.crd.york.ac.uk/PROSPERO) holds the registration record of this systematic review.

Results from the systematic review will be disseminated via peer reviewed publications and relevant academic conferences. Findings may also be published in relevant professional and third sector newsletters and magazines as appropriate. Data will be used to inform future research and guideline development.

### Outcome of systematic review

A comprehensive understanding of how grounded theory has been used in relation to the study of hearing loss will be identified. Overall, a set of over-arching themes arising from the body of qualitative research that focused on adults’ experiences of hearing loss will be established.

## Data Availability

N/A

## Summary

This systematic review will be the first to establish how grounded theory has been used to investigate hearing loss. A critical appraisal of all hearing loss studies that have investigated acquired hearing loss within adult populations using grounded theory will be performed. This novel systematic review will aid in implementing more precise applications of grounded theory in future research. This will also enable more refined understandings of the experiences of hearing loss to be established and facilitate better care for those who have hearing loss.

## Contributors

YA is the guarantor of the review (CRD42019134197). YA led on the development of the review protocol and drafted the manuscript. YA, NW, DC, HH and DH contributed to the development of the eligibility criteria, and selection process. NW, DC, HH, and DH all read drafts of the manuscript, provided feedback, and approved the final manuscript.

## Exclusive license

The corresponding Author, Yasmin Ali, has the right to grant on behalf of all authors and does grant on behalf of all authors, an exclusive licence (or non-exclusive for government employees) on a worldwide basis to the BMJ Publishing Group Ltd to permit this article (if accepted) to be published in BMJ editions and any other BMJPGL products and sublicenses such use and exploit all subsidiary rights, as set out in our licence.

## Disclaimer

The views expressed are those of the authors and not necessarily those of the NHS, the NIHR, or the Department of Health and Social Care.

## Transparency declaration

Yasmin Ali affirms that the manuscript is an honest, accurate, and transparent account of the study being reported; that no important aspects of the study have been omitted; and that any discrepancies from the study as planned have been explained.

## Patient and public involvement

There was no patient and public involvement due to the methodological nature of this study.

## Provenance and peer review

Not commissioned; externally peer reviewed.

## Data sharing statement

All data collected according to the data items will be available on request if not included in the final published systematic review article.

## References

1 Alshuaib, W. B., Al-Kandari, J. M. & Hasan, S. M. Update On Hearing Loss. InTech (2015). 2 World Health Organization. Addressing the rising prevalence of hearing loss. Report No. 9241550260, (World Health Organization, 2018).

2 Action On Hearing Loss. Hearing Matters Report. (2015). (Available at: https://www.actiononhearingloss.org.uk/-/media/ahl/documents/research-and-policy/reports/hearing-matters-report.pdf

3 World Health Organization. Global costs of unaddressed hearing loss and cost-effectiveness of interventions: a WHO report, 2017. Report No. 9241512040, (World Health Organization, 2017).

4 Archbold, S., Lamb, B., O’Neill, C. & Atkins, J. The real cost of adult hearing loss: reducing its impact by increasing access to the latest hearing technologies. The Ear Foundation, EarFoundation (Erişim: 12.08. 2017) (2014).

5 Dobie, R. A., Van Hemel, S. & Council, N. R. Basics of Sound, the Ear, and Hearing. (2004).

6 Dalton, D. S. et al. The impact of hearing loss on quality of life in older adults. Gerontologist 43, 661–668 (2003).

7 Knutson, J. F. & Lansing, C. R. The relationship between communication problems and psychological difficulties in persons with profound acquired hearing loss. Journal of Speech and Hearing Disorders 55, 656–664 (1990).

8 Demorest, M. E. & Erdman, S. A. Development of the communication profile for the hearing impaired. Journal of Speech and Hearing Disorders 52, 129–143 (1987).

9 Weinstein, B. E. & Ventry, I. M. Hearing impairment and social isolation in the elderly. J Speech Hear Res 25, 593–599 (1982).

10 Mick, P., Kawachi, I. & Lin, F. R. The Association between Hearing Loss and Social Isolation in Older Adults. Otolaryng Head Neck 150, 378–384, doi:10.1177/0194599813518021 (2014).

11 Heffernan, E., Coulson, N. S. & Ferguson, M. A. Development of the Social Participation Restrictions Questionnaire (SPaRQ) through consultation with adults with hearing loss, researchers, and clinicians: a content evaluation study. International Journal of Audiology 57, 791–799, doi:10.1080/14992027.2018.1483585 (2018).

12 Arlinger, S. Negative consequences of uncorrected hearing loss-a review. International journal of audiology 42, 2S17–12S20 (2003).

13 Gomaa, M. A. M., Elmagd, M. H. A., Elbadry, M. M. & Kader, R. M. A. J. E. A. o. O.-R.-L. Depression, Anxiety and Stress Scale in patients with tinnitus and hearing loss. 271, 2177–2184 (2014).

14 Kvam, M. H., Loeb, M., Tambs, K. J. T. J. o. d. s. & education, d. Mental health in deaf adults: symptoms of anxiety and depression among hearing and deaf individuals. 12, 1–7 (2007).

15 Golub, J. S. et al. Association of Audiometric Age-Related Hearing Loss With Depressive Symptoms Among Hispanic Individuals. Jama Otolaryngol 145, 132–139, doi:10.1001/jamaoto.2018.3270 (2019).

16 Thomas, A. J. Acquired hearing loss: Psychological and psychosocial implications. (Academic Press, 1984).

17 Barker, A. B., Leighton, P. & Ferguson, M. A. Coping together with hearing loss: a qualitative meta-synthesis of the psychosocial experiences of people with hearing loss and their communication partners. International Journal of Audiology 56, 297–305, doi:10.1080/14992027.2017.1286695 (2017).

18 Hallberg, L. R. & Barrenas, M. L. Living with a male with noise-induced hearing loss: experiences from the perspective of spouses. Br J Audiol 27, 255–261 (1993).

19 Hetu, R., Jones, L. & Getty, L. The impact of acquired hearing impairment on intimate relationships: implications for rehabilitation. Audiology 32, 363–381 (1993).

20 Heffernan, E., Coulson, N. S., Henshaw, H., Barry, J. G. & Ferguson, M. A. Understanding the psychosocial experiences of adults with mild-moderate hearing loss: An application of Leventhal’s self-regulatory model. International Journal of Audiology 55, S3–S12, doi:10.3109/14992027.2015.1117663 (2016).

21 Coker, A. L., Smith, P. H., Bethea, L., King, M. R. & McKeown, R. E. Physical health consequences of physical and psychological intimate partner violence. Arch Fam Med 9, 451–457, doi:DOI 10.1001/archfami.9.5.451 (2000).

22 Bunne, M. Qualitative research methods in otorhinolaryngology. Int J Pediatr Otorhinolaryngol 51, 1–10 (1999).

23 Knudsen, L. V. et al. Conducting qualitative research in audiology: A tutorial. 51, 83–92 (2012).

24 Mugenda, O. M. Research methods: Quantitative and qualitative approaches. (African Centre for Technology Studies, 1999).

25 Creswell, J. W. & Poth, C. N. Qualitative inquiry and research design: Choosing among five approaches. (Sage publications, 2017).

26 Glaser, B. G., Strauss, A. L. & Strutzel, E. The discovery of grounded theory; strategies for qualitative research. Nursing research 17, 364 (1968).

27 Charmaz, K. Constructing grounded theory: A practical guide through qualitative analysis. (Sage, 2006).

28 Glaser, B. G. & Strauss, A. L. Discovery of grounded theory: Strategies for qualitative research. (Routledge, 2017).

29 Fassinger, R. E. Paradigms, praxis, problems, and promise: Grounded theory in counseling psychology research. Journal of counseling psychology 52, 156 (2005).

30 Strauss, A. & Corbin, J. M. Grounded theory in practice. (Sage, 1997).

31 Weed, M. Research quality considerations for grounded theory research in sport & exercise psychology. Psychology of sport and exercise 10, 502–510 (2009).

32 Lazenbatt, A. & Elliott, N. How to recognise a’quality’grounded theory research study. Australian Journal of Advanced Nursing, The 22, 48 (2005).

33 Ali, N., May, S. & Grafton, K. A systematic review of grounded theory studies in physiotherapy. Physiotherapy theory and practice, 1-31 (2018).

34 Hutchison, A. J., Johnston, L. & Breckon, J. Grounded theory-based research within exercise psychology: A critical review. Qualitative Research in Psychology 8, 247–272 (2011).

35 Douglas, D. Grounded theories of management: A methodological review. Management Research News 26, 44–52 (2003).

36 da Silva Barreto, M., Garcia-Vivar, C. & Marcon, S. S. Methodological quality of Grounded Theory research with families living with chronic illness. International journal of Africa nursing sciences 8, 14–22 (2018).

37 Charmaz, K. ‘Discovering’chronic illness: using grounded theory. Soc Sci Med 30, 1161–1172 (1990).

38 Baker, C. & Stern, P. N. Finding meaning in chronic illness as the key to self-care. Canadian Journal of Nursing Research Archive 25 (1993).

39 Conrad, P. Qualitative research on chronic illness: a commentary on method and conceptual development. Soc Sci Med 30, 1257–1263 (1990).

40 Glaser, B. G. & Strauss, A. L. Awareness of dying. (Routledge, 2017).

41 Corbin, J. & Strauss, A. Managing chronic illness at home: three lines of work. Qualitative sociology 8, 224–247 (1985).

42 Belgrave, L. L. & Charmaz, K. in The Social Construction of Death 34-51 (Springer, 2014).

43 Charmaz, K. Loss of self: a fundamental form of suffering in the chronically ill. Sociology of health & illness 5, 168–195 (1983).

44 Charmaz, K. Grounded theory as an emergent method. Handbook of emergent methods 155, 172 (2008).

45 Bernell, S. & Howard, S. W. Use your words carefully: what is a chronic disease? Frontiers in public health 4, 159 (2016).

46 World Health Organization. Preventing chronic diseases: a vital investment. Report No. 9241563001, (World Health Organization, Public Health Agency of Canada, Canada. Public Health Agency of Canada). (2005).

47 Scinicariello, F., Carroll, Y., Eichwald, J., Decker, J. & Breysse, P. N. Association of Obesity with Hearing Impairment in Adolescents. Sci Rep-Uk 9, 1877 (2019).

48 Blackwell, D. L., Lucas, J. W. & Clarke, T. C. Summary health statistics for US adults: national health interview survey, 2012. Vital and health statistics. Series 10, Data from the National Health Survey, 1-161 (2014).

49 Vos, T. et al. Global, regional, and national incidence, prevalence, and years lived with disability for 310 diseases and injuries, 1990–2015: a systematic analysis for the Global Burden of Disease Study 2015. The Lancet 388, 1545–1602 (2016).

50 Ferguson, M. A. et al. Hearing aids for mild to moderate hearing loss in adults. Cochrane Db Syst Rev, doi:10.1002/14651858.CD012023.pub2 (2017).

51 Maidment, D. W., Barker, A. B., Xia, J. & Ferguson, M. A. Effectiveness of alternative listening devices to conventional hearing aids for adults with hearing loss: a systematic review protocol. BMJ open 6, e011683 (2016).

52 Mamo, S. K. et al. Hearing loss treatment in older adults with cognitive impairment: A systematic review. Journal of Speech, Language, and Hearing Research 61, 2589–2603 (2018).

53 Henshaw, H. & Ferguson, M. A. Efficacy of individual computer-based auditory training for people with hearing loss: A systematic review of the evidence. PloS one 8, e62836 (2013).

54 Yu, H. & Li, H. Association of vertigo with hearing outcomes in patients with sudden sensorineural hearing loss: a systematic review and meta-analysis. JAMA otolaryngology– head & neck surgery 144, 677–683 (2018).

55 Nordvik, Ø. et al. Generic quality of life in persons with hearing loss: a systematic literature review. BMC Ear, Nose and Throat Disorders 18, 1 (2018).

56 Lawrence, B. J. et al. Hearing loss and depression in older adults: A systematic review and meta-analysis. The Gerontologist (2019).

57 Moher, D. et al. Preferred reporting items for systematic review and meta-analysis protocols (PRISMA-P) 2015 statement. Syst Rev 4, doi:10.1186/2046-4053-4-1 (2015).

58 Shamseer, L. et al. Preferred reporting items for systematic review and meta-analysis protocols (PRISMA-P) 2015: elaboration and explanation. Bmj-Brit Med J 349, doi:10.1136/bmj.g7647 (2015).

59 Munn, Z., Stern, C., Aromataris, E., Lockwood, C. & Jordan, Z. What kind of systematic review should I conduct? A proposed typology and guidance for systematic reviewers in the medical and health sciences. Bmc Med Res Methodol 18, doi:10.1186/s12874-017-0468-4 (2018).

60 Clarke, M., Oxman, A., Paulsen, E., Higgins, J. & Green, S. Appendix A: Guide to the contents of a Cochrane Methodology protocol and review. Cochrane Handbook for Systematic Reviews of Interventions (2009).

61 McIlroy, G. & Storbeck, C. Development of deaf identity: an ethnographic study. J Deaf Stud Deaf Educ 16, 494–511, doi:10.1093/deafed/enr017 (2011).

62 Chapman, M. & Dammeyer, J. The Significance of Deaf Identity for Psychological Well-Being. J Deaf Stud Deaf Educ, doi:10.1093/deafed/enw078 (2016).

63 Leigh, I., Marcus, A., Dobosh, P. & Allen, T. Deaf/hearing cultural identity paradigms: modification of the deaf identity development scale. J Deaf Stud Deaf Educ 3, 329–338, doi:10.1093/oxfordjournals.deafed.a014360 (1998).

64 Audiology, B. S. o. Recommended procedure: pure-tone air-conduction and bone- conduction threshold audiometry with and without masking. United Kingdom; (2017).

65 Berthelsen, C. B., Grimshaw-Aagaard, S. L. S. & Hansen, C. Developing a Guideline for Reporting and Evaluating Grounded Theory Research Studies (GUREGT). International Journal of Health Sciences 6, 64–76 (2018).

66 Braun, V. & Clarke, V. What can “thematic analysis” offer health and wellbeing researchers? Int J Qual Stud Heal 9, doi:10.3402/qhw.v9.26152 (2014).

67 Clarke, V. & Braun, V. Thematic analysis. J Posit Psychol 12, 297–298, doi:10.1080/17439760.2016.1262613 (2017).

68 Armstrong, D., Gosling, A., Weinman, J. & Marteau, T. The place of inter-rater reliability in qualitative research: an empirical study. Sociology 31, 597–606 (1997).

69 McAlister, A. et al. Qualitative coding: An approach to assess inter-rater reliability. In ASEE Annual Conference & Exposition, Columbus, Ohio. https://peer.asee.org/28777. (2017).

